# Adverse Drug Reaction Reporting Among Herbal Practitioners: A Mixed-Methods Study of Reporting Practices, Associated Factors, and Pharmacovigilance System Barriers in the Greater Accra Region, Ghana

**DOI:** 10.64898/2026.10.06.26364851

**Authors:** Harriet Fianko, Joseph Sam, Kwabena Frimpong-Manso Opuni, Seth Kwaku Seaneke, Colette Santah, Paul Botwe, Francisca Ziga, George Tsey Sabblah, Harriet Affran Bonful

## Abstract

**Background:** Herbal medicinal products are widely used in Ghana; however, adverse drug reaction (ADR) reporting associated with these products remains limited, posing challenges to pharmacovigilance and medicine safety surveillance. This study assessed trends in herbal medicine-related ADR reporting, determined the prevalence and factors associated with ADR reporting among herbal practitioners, and explored barriers to ADR reporting in Ghana.

**Methods:** A mixed-methods study involving secondary pharmacovigilance data review, an analytic cross-sectional survey, and qualitative in-depth interviews was conducted. Secondary data on herbal medicine-related ADR reports submitted to the Ghana Food and Drugs Authority (FDA) between 2014 and 2024 were reviewed descriptively. The cross-sectional survey involved 271 herbal practitioners selected through multi-stage probability sampling across seven districts in the Greater Accra Region. Quantitative data were analysed using descriptive statistics and logistic regression in IBM SPSS Statistics version 26. The qualitative component involved 15 purposively selected participants, and thematic analysis was conducted using MAXQDA 2020.

**Results:** A total of 117 herbal medicine-related ADR reports met the inclusion criteria and were analysed from the Ghana FDA pharmacovigilance database between 2014 and 2024. Among 271 surveyed herbal practitioners, 11.8% (95% CI: 8.0–15.6) reported submitting at least one ADR report during the preceding 12 months. In multivariable analysis, lack of ADR reporting forms or job aids (aOR = 0.11, 95% CI: 0.04–0.31) and lack of exposure to Ghana FDA ADR reporting guidelines (aOR = 0.09, 95% CI: 0.03–0.32) were independently associated with lower odds of ADR reporting. Qualitative findings identified causal uncertainty, inadequate training, weak reporting systems, commercial interests, and limited regulatory support as major barriers to reporting.

**Conclusion:** ADR reporting associated with herbal medicinal products was limited within the national pharmacovigilance system and among surveyed herbal practitioners. System-level barriers, particularly limited access to reporting tools and FDA reporting guidance, were associated with lower reporting. Strengthening pharmacovigilance infrastructure, improving practitioner training, and increasing access to reporting tools may improve herbal medicine safety surveillance.

## Introduction

Herbal medicinal products are widely used globally and remain an important component of healthcare delivery, particularly in low-and middle-income countries (LMICs) where access to conventional healthcare may be limited. The World Health Organization (WHO) estimates that a substantial proportion of populations in Africa relies on traditional and herbal medicine for primary healthcare needs^1^. In Ghana, the use of herbal medicine remains widespread due to cultural acceptance, accessibility, affordability, and perceived effectiveness^2^.

Despite their widespread use, herbal medicinal products are associated with important safety concerns. Adverse drug reactions (ADRs) linked to herbal medicines, including hepatotoxicity, nephrotoxicity, cardiotoxicity, allergic reactions, and herb–drug interactions, have been documented in previous studies^3^. The increasing availability of herbal products through formal and informal markets, coupled with self-medication practices and inconsistent regulation, has heightened concerns regarding the safety monitoring of these products.

Pharmacovigilance systems are important for identifying and preventing ADRs associated with medicinal products, including herbal medicines^4^. Effective ADR reporting is particularly important for herbal medicines because many products contain multiple bioactive constituents, may vary in composition, and are frequently used concurrently with conventional medicines^3, 4^.

However, underreporting of ADRs remains a major global pharmacovigilance challenge and appears to be particularly pronounced in the herbal medicine sector. Previous studies among healthcare professionals have identified several barriers to ADR reporting, including limited pharmacovigilance knowledge, uncertainty in establishing causality, inadequate reporting systems, lack of training, and weak institutional support^5, 6^. In many low-resource settings, herbal medicinal products pharmacovigilance remains weakly institutionalized, resulting in limited reporting and inadequate safety data.

Following the Ministry of Health’s initiative to integrate Traditional, Complementary and Alternative Medicine (TCAM), particularly herbal medicine, into Ghana’s healthcare system, the National Health Policy was revised to promote collaboration between traditional and conventional healthcare services^6^. This policy direction facilitated the formal integration of herbal medicine into selected public health facilities in 2012 and subsequent efforts to strengthen herbal medicine practice within the national health system^7^. As a result, herbal medicine has increasingly become part of formal healthcare delivery in Ghana, creating opportunities for collaboration between herbal practitioners and orthodox healthcare professionals although challenges remain in areas such as referral systems, communication, and pharmacovigilance^8^. Consequently, effective ADR reporting and safety monitoring have become increasingly important to ensure the safe integration and use of herbal medicinal products within the healthcare system.

In Ghana, the Food and Drugs Authority (FDA) coordinates national pharmacovigilance activities, including ADR reporting for herbal medicinal products. Although pharmacovigilance systems have expanded over time, reporting of herbal medicinal products-related ADRs remains relatively low compared with conventional medicines. Existing studies in Ghana have largely focused on healthcare professionals such as physicians, pharmacists, and nurses, with limited attention given to herbal practitioners despite their central role in herbal medicine use and distribution^2, 9, 10^. Furthermore, evidence on factors influencing ADR reporting among herbal practitioners and the systemic barriers affecting reporting practices remains limited^3^. This study addresses this gap by combining national pharmacovigilance data with quantitative and qualitative evidence to examine ADR reporting practices and the individual and system-level factors influencing reporting behaviour among herbal practitioners in Ghana.

Understanding the factors associated with ADR reporting among herbal practitioners is important for strengthening herbal medicinal products pharmacovigilance systems and improving medicine safety surveillance in Ghana. Therefore, this study aimed to assess trends in herbal medicine-related ADR reporting, determine the prevalence and factors associated with ADR reporting among practitioners, and explore barriers to reporting in the Greater Accra Region of Ghana.

## Methods

### Study Design

This study employed a mixed-methods design combining descriptive analysis of national adverse drug reaction (ADR) reports related to herbal medicinal products to assess reporting trends, a cross-sectional survey to determine the prevalence and factors associated with ADR reporting, and in-depth interviews to explore barriers and contextual influences on reporting practices. Triangulation of findings from these complementary data sources provided a more comprehensive understanding of ADR reporting among herbal practitioners in Ghana.

### Study Setting

Herbal medicine is an important component of healthcare delivery in Ghana and is regulated by the Traditional Medicine Practice Council (TMPC) and the Food and Drugs Authority (FDA). At the national level, over 6,000 practitioners, more than 5,000 premises, and about 20 training institutions are registered with the TMPC^11^. Herbal practitioners operate through herbal clinics, herbal pharmacies, and community-based practices, where they provide consultation and treatment services using approved herbal medicinal products for a variety of acute and chronic health conditions. The Greater Accra Region hosts a substantial proportion of these facilities and practitioners, making it an appropriate setting for assessing adverse drug reaction (ADR) reporting practices among herbal practitioners^12^.

### Data collection

Data were collected in three phases. Phase 1 involved the secondary data analysis of ADR reports related to herbal medicinal products obtained from the Ghana Food and Drugs Authority national pharmacovigilance database. The second phase covered the analytical cross-sectional study among herbal practitioners, and the last phase entailed the qualitative study involving in-depth interviews with selected practitioners.

### Phase 1-Secondary Pharmacovigilance Data analysis

Secondary data were obtained from the national pharmacovigilance database of the Ghana Food and Drugs Authority (FDA), on 8^th^ September 2025. The data base contains ADR reports submitted through the national reporting system. It comprised ADR reports related to herbal medicinal products recorded between 2014 and 2024. Variables extracted included reporting year, patient demographics, seriousness of reaction, system organ class (SOC), reporter category, geographical region, and patient outcome. A total of 122 reports were initially identified. One duplicate report and four reports with incomplete information on reporting year were excluded, leaving 117 reports that met the inclusion criteria and were analysed.

## Phase 2-Analytic cross-sectional survey among Herbal Practitioners

### Study Design and Population

A cross-sectional survey was conducted among herbal practitioners working in herbal clinics and herbal pharmacies within the seven districts (Adenta Municipal, La Nkwantanang Madina Municipal, Ablekuma Central Municipal, Ga East Municipal, Ga West Municipal, Ayawaso Central Municipal, and Ayawaso West Municipal) in the Greater Accra Region.

In this study, the term “herbal practitioners” was used as an umbrella term to refer to individuals involved in herbal medicine practice, including herbal /traditional healers and community herbal retailers or sellers. Those who experienced a language barrier with English were excluded.

### Sampling and Sample Size

A multistage probability sampling approach was used. First, seven districts in the Greater Accra Region were selected using simple random sampling. Second, a sampling frame of registered herbal practitioners within the selected districts was obtained from the Traditional Medicine Practice Council (TMPC) registry. The required sample was proportionately allocated across the seven districts based on the number of registered practitioners in each district. Finally, eligible practitioners within each district were assigned unique identification numbers and selected using computer-generated random numbers. A total of 271 herbal practitioners participated in the survey.

### Study Variables

The outcome variable was self-reported ADR reporting related to herbal medicinal products within the previous 12 months (“Yes” or “No”).

Independent variables were grouped into socio-demographic, individual-level, and system-level factors based on previous pharmacovigilance literature^5, 6^. Socio-demographic variables included age, educational level, profession, work experience, and workplace setting. Individual-level variables included ADR knowledge level and number of suspected ADR cases encountered within the previous 12 months. System-level variables included availability of ADR reporting tools, exposure to Ghana FDA ADR reporting guidelines, and formal ADR reporting training.

## Data Collection

Data were collected using a structured questionnaire developed by the researchers based on the study objectives and a review of the pharmacovigilance literature on adverse drug reaction (ADR) reporting, pharmacovigilance knowledge, training, reporting systems, and barriers to reporting^5, 13^. The questionnaire comprised sections on sociodemographic characteristics, ADR reporting practices, knowledge of pharmacovigilance and ADR reporting, training and reporting systems, and perceived barriers to reporting. The questionnaire was pretested before data collection.

## Data Analysis

Secondary pharmacovigilance data were analysed descriptively to summarize reporting trends and characteristics of herbal medicine-related ADR reports.

Data obtained from the cross-sectional study data were analysed using IBM SPSS Statistics version 26. Descriptive statistics summarized participant characteristics and ADR reporting prevalence. Bivariate associations between explanatory variables and ADR reporting were assessed using chi-square tests. Variables that were statistically significant at the bivariate level (p < 0.05) or considered theoretically relevant were included in the multivariable logistic regression model. The final model was specified parsimoniously, considering the limited number of ADR reporting events and potential conceptual overlap among explanatory variables. Adjusted odds ratios (aORs) with 95% confidence intervals (CIs) were reported. All statistical tests were two-tailed, with p < 0.05 considered statistically significant.

## Qualitative Component

### Study Design and Sampling

The qualitative component involved in-depth interviews with purposively selected herbal practitioners with at least two years of professional experience. Purposive sampling was used to obtain information-rich narratives related to ADR reporting experiences and barriers. A total of 15 interviews were conducted until thematic sufficiency was achieved.

### Data Collection and Analysis

Interviews were conducted in English using a semi-structured interview guide and were audio-recorded with participant consent. Audio recordings were transcribed verbatim and analysed thematically using both inductive and deductive approaches. Transcripts were repeatedly read to achieve familiarization with the data and to contextualize participants’ account. Coding and data management were then conducted using MAXQDA version 2020. Codes representing meaningful units of text were generated and compared across transcripts to identify patterns and variations. Related codes were grouped into categories and refined into themes and subthemes. Themes were then reviewed to ensure coherence and clear distinctions between categories. The final themes and subthemes formed the basis for organising the qualitative results and interpretations of the findings in relation to existing literature.

## Ethical Considerations

Ethical approval was obtained from the Noguchi Memorial Institute for Medical Research Institutional Review Board (NMIMR-IRB CPN 020/25-26). Written informed consent was obtained from all participants prior to data collection. Confidentiality and anonymity were maintained throughout the study.

## Results

### Secondary Pharmacovigilance Data on Herbal Medicine-Related ADR Reports

A total of 122 adverse drug reaction (ADR) reports related to herbal medicinal products were identified in the Ghana Food and Drugs Authority (FDA) pharmacovigilance database between 2014 and 2024. One duplicate report and four reports with incomplete information on reporting year were excluded, leaving 117 reports for analysis. The characteristics of the 117 herbal medicine-related ADR reports are presented in Table 1.

**Table 1.**
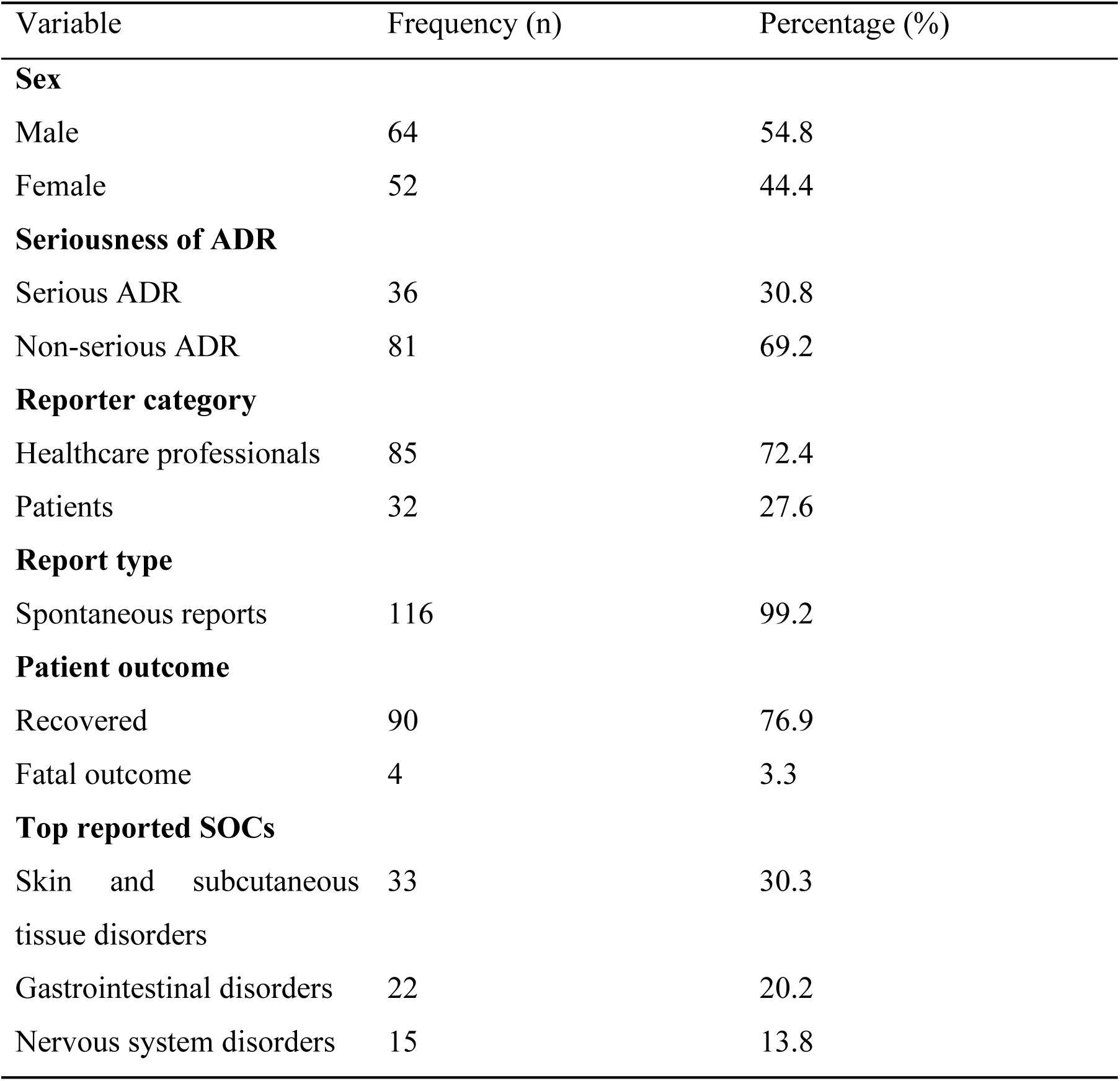
Characteristics of Herbal Medicine-Related ADR Reports Submitted to the Ghana FDA, 2014–2024 (n = 117)

More than half of the ADR reports involved male patients (64/117; 54.8%), while females accounted for 44.4% (52/117). Most ADRs were classified as non-serious (81/117; 69.2%), whereas 30.8% (36/117) were categorized as serious. Healthcare professionals submitted the majority of reports (85/117; 72.4%), while patient reporting constituted 27.6% (32/117). Nearly all reports were spontaneous reports (116/117; 99.2%).

### Trends in Herbal Medicine-Related ADR Reports

The number of herbal medicine-related ADR reports fluctuated across the study period without a consistent upward trend. Reporting peaked in 2018 (25 reports) and 2016 (23 reports), followed by a gradual decline in subsequent years. The lowest number of reports was recorded in 2014, with only two reports documented. Reporting remained relatively low in recent years, with seven reports each recorded in 2023 and 2024. The annual trend in herbal medicine-related ADR reports submitted to the Ghana FDA from 2014 to 2024 is shown in Figure 1.

**Figure 1:**
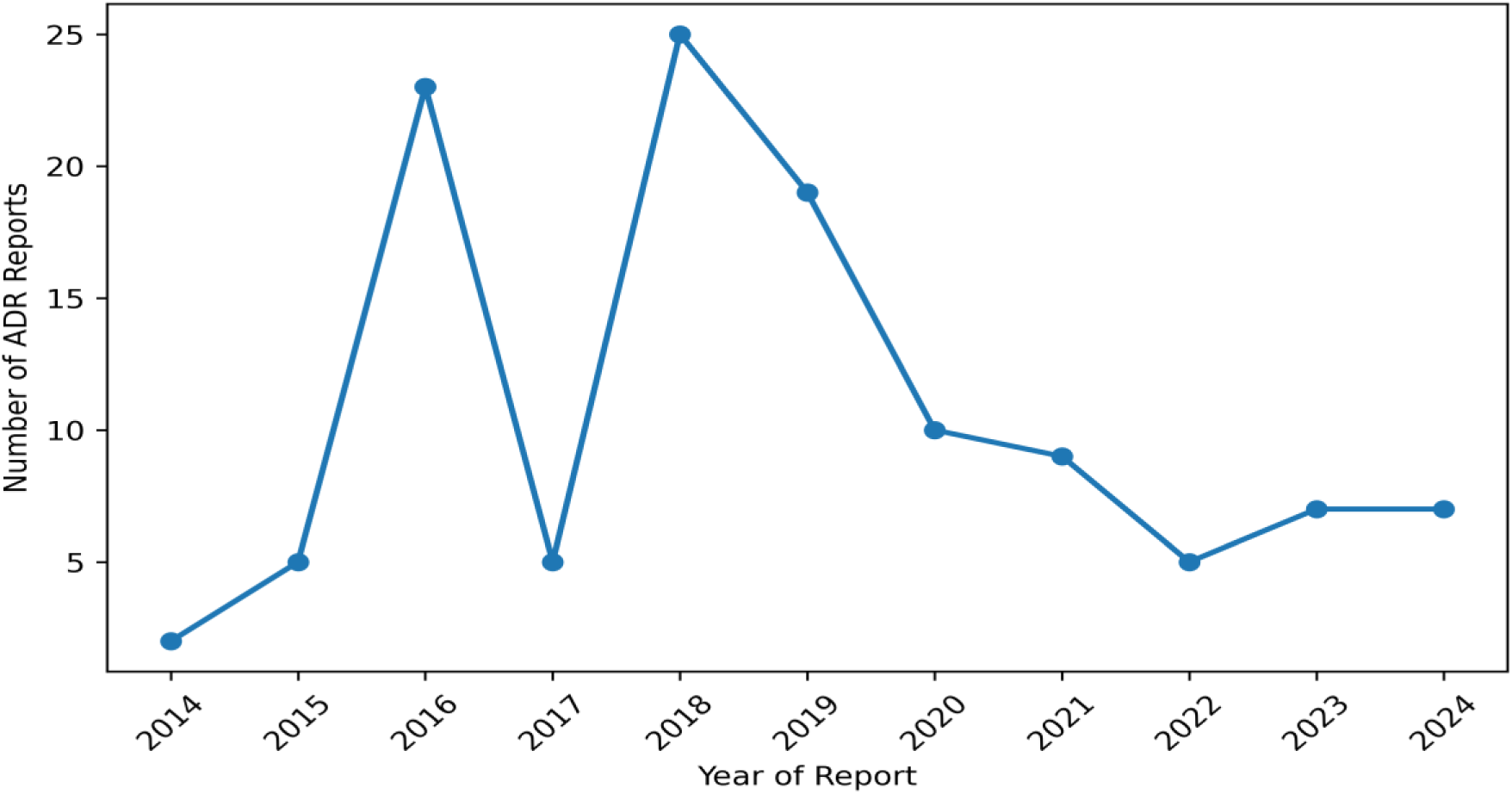
Annual Trends in Herbal Medicine-Related ADR Reports Submitted to the Ghana FDA, 2014–2024

### Regional Distribution and Patient Outcomes

Greater Accra recorded the highest number of ADR reports (43/117; 36.8%) and was the only region that consistently submitted reports throughout the study period. The Volta Region accounted for 12.8% (15/117) of reports, while Ashanti and Central Regions each contributed 8.6% (10/117). Northern, Upper West, and Upper East regions recorded comparatively fewer reports which are 6 (5.1%), 5 (4.3%), 4 (3.4%) respectively. Most patients recovered following the reported ADRs (90/117; 76.9%), while 3.3% (4/117) of reports resulted in fatal outcomes. Fatal cases were recorded in 2015, 2016, and 2019. Fatal outcomes were recorded among reports classified as serious ADRs.

### System Organ Class Distribution

The most commonly reported system organ classes (SOCs) were Skin and Subcutaneous Tissue Disorders (30.3%), Gastrointestinal Disorders (20.2%), and Nervous System Disorders (13.8%). Less frequently reported SOCs included Cardiac Disorders, Eye Disorders, and Vascular Disorders. The distribution of the five most frequently reported System Organ Classes (SOCs) is shown in Figure 2

**Figure 2:**
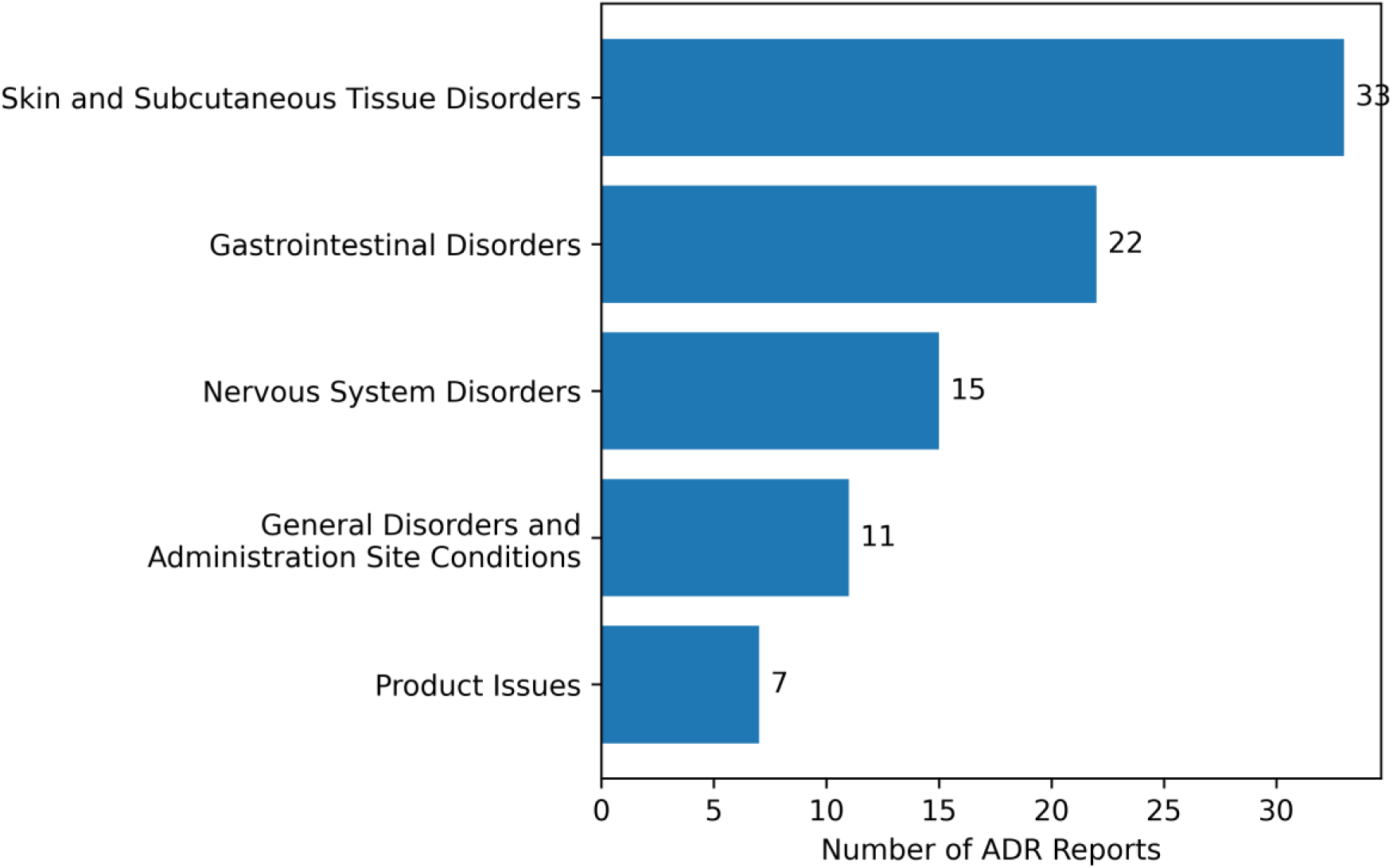
Top Five Reported Herbal Medicine-Related Adverse Drug Reaction System Organ Classes (SOCs), Ghana FDA Database, 2014–2024

### Socio-demographic Characteristics of Survey Participants

A total of 271 herbal practitioners participated in the quantitative survey. Most respondents were aged 35–44 years (103/271; 38.0%), followed by those aged 45–54 years (67/271; 24.7%). Only 2.6% (7/271) of participants were below 25 years of age. Regarding educational level, 36.5% (99/271) had tertiary education, while 30.6% (83/271) had completed senior high school. Participants with herbal medicine degrees or traditional medicine training accounted for 11.8% (32/271) of respondents. Community herbal retailers or sellers constituted the largest professional category (137/271; 50.6%), followed by herbal practitioners or traditional healers (109/271; 40.2%). Most respondents had between one and five years of work experience (102/271; 37.6%), while 26.2% (71/271) had more than ten years of experience. Herbal pharmacy or retail settings represented the most common workplace setting (151/271; 55.7%). The socio-demographic characteristics of the herbal practitioners who participated in the study are presented in Table 2.

**Table 2.**
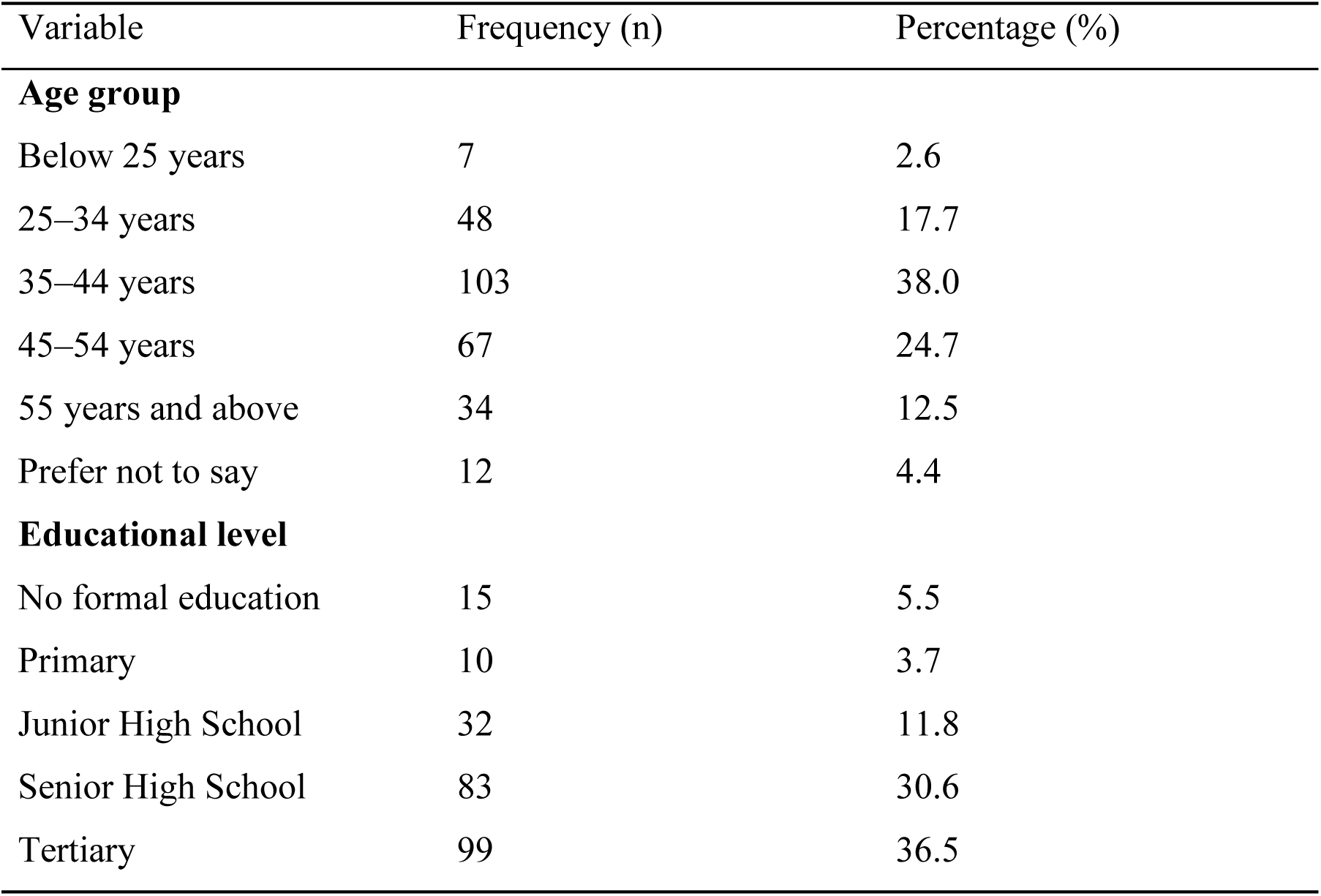
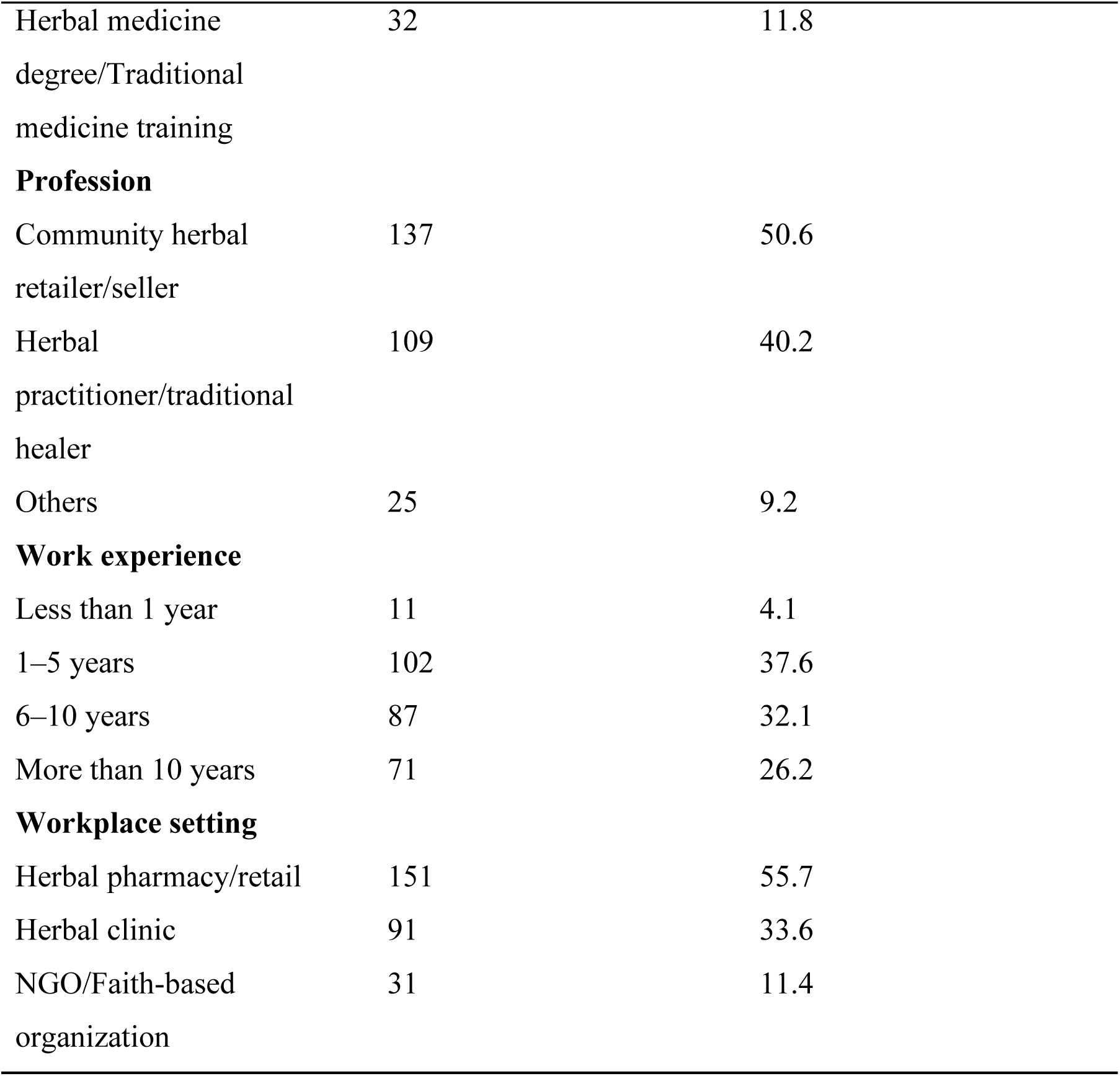
Socio-demographic Characteristics of Herbal Practitioners Participating in the Study (n = 271)

### Knowledge, Training, and System-Related Characteristics

Regarding knowledge, training, and system-related characteristics, 50.9% (138/271) of herbal practitioners had low ADR knowledge, while 49.1% (133/271) had high ADR knowledge. Only 14.4% (39/271) had received formal ADR training within the previous two years, whereas 85.6% (232/271) had not. In addition, 26.9% (73/271) reported having seen the Ghana FDA ADR reporting guideline or reporting form, while 73.1% (198/271) had not. The knowledge, training, and system-related characteristics of the herbal practitioners are presented in Table 3.

**Table 3.**
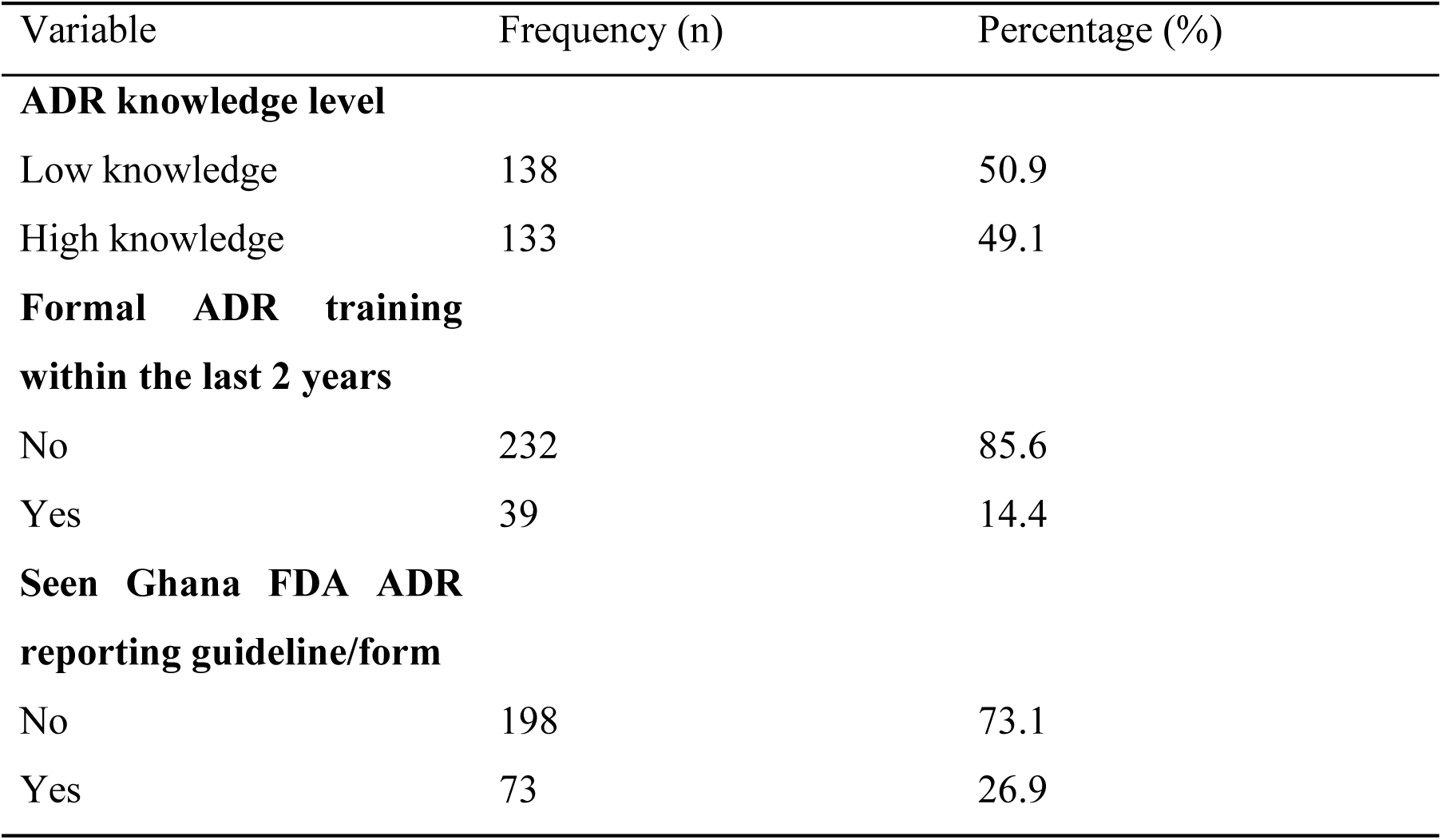
Knowledge, Training, and System-Related Characteristics of Herbal Practitioners.

### Prevalence of ADR Reporting Among Herbal Practitioners

Out of the 271 respondents included in the study, only 32 (11.8%) reported having submitted at least one ADR report related to herbal medicinal products within the previous 12 months, while 239 (88.2%) had not submitted any report. The estimated 12-month prevalence of ADR reporting among herbal practitioners was 11.8% (95% CI: 8.0–15.6).

### Factors Associated with ADR Reporting Among Herbal Practitioners

Bivariate analyses were conducted to assess associations between selected socio-demographic, individual-level, and system-level factors and ADR reporting among herbal practitioners.

### Socio-demographic Factors

Bivariate analysis showed significant associations between several socio-demographic characteristics and ADR reporting. Age group was significantly associated with ADR reporting (χ² = 13.70, p = 0.018), with respondents aged 35–44 years accounting for the largest proportion of ADR reports. Educational level was also significantly associated with ADR reporting (χ² = 46.06, p < 0.001), particularly among respondents with herbal medicine degrees or traditional medicine training. Profession was significantly associated with ADR reporting (χ² = 10.51, p = 0.005), with herbal practitioners or traditional healers accounting for a greater proportion of those who reported ADRs than community herbal retailers (Table 4). However, work experience was not significantly associated with ADR reporting (p = 0.331).

**Table 4.** Bivariate Analysis of Factors Associated with ADR Reporting Among Herbal Practitioners (n = 271)

| Factor | $\chi^2$ (df) | p-value |
| --- | --- | --- |
| **Socio-demographic factors** |  |  |
| Age group | 13.70 (5) | 0.018 |
| Educational level | 46.06 (5) | <0.001 |
| Profession | 10.51 (2) | 0.005 |
| Work experience | 3.42 (3) | 0.331 |
| **Individual-level factors** |  |  |
| ADR knowledge level | 15.03 (1) | <0.001 |
| Formal ADR training | 5.56 (1) | 0.018 |
| Number of ADR cases encountered | 97.06 (3) | <0.001 |
| **System-level factors** |  |  |
| Availability of ADR forms/posters/job aids | 72.12 (2) | <0.001 |
| Seen Ghana FDA ADR reporting guideline/form | 54.39 (1) | <0.001 |
| Herbal clinic setting | 8.36 (1) | 0.004 |
| Herbal pharmacy/retail setting | 13.88 (1) | <0.001 |
| NGO/faith-based setting | 1.91 (1) | 0.166 |

### Individual-Level Factors

ADR knowledge level was significantly associated with ADR reporting (χ² = 15.03, p < 0.001), with respondents who demonstrated higher knowledge more likely to report ADRs (Table 4). Formal ADR training within the previous two years was also associated with ADR reporting (χ² = 5.56, p = 0.018). In addition, the number of suspected ADR cases encountered within the previous 12 months showed a strong association with reporting behaviour (χ² = 97.06, p < 0.001).

### System-Level Factors

Several system-level factors were significantly associated with ADR reporting (Table 4). Availability of ADR reporting forms, posters, or job aids in the facility was strongly associated with ADR reporting (χ² = 72.12, p < 0.001). Similarly, respondents who had seen the Ghana FDA ADR reporting guideline or reporting form were significantly more likely to report ADRs (χ² = 54.39, p < 0.001). Workplace setting was also associated with ADR reporting, with respondents working in herbal clinics more likely to report ADRs (χ² = 8.36, p = 0.004), while those working in herbal pharmacy or retail settings were less likely to report ADRs (χ² = 13.88, p < 0.001).

The bivariate associations between these factors and ADR reporting are presented in Table 4.

### Multivariable Logistic Regression Analysis

Table 5 shows that in the crude logistic regression analysis, respondents with low ADR knowledge had significantly lower odds of reporting ADRs compared with those with high knowledge (cOR = 0.19, 95% CI: 0.07–0.47). Similarly, absence of ADR reporting forms or job aids in the facility (cOR = 0.05, 95% CI: 0.02–0.11) and lack of exposure to the Ghana FDA ADR reporting guideline (cOR = 0.06, 95% CI: 0.02–0.15) were associated with substantially lower odds of ADR reporting.

**Table 5.**
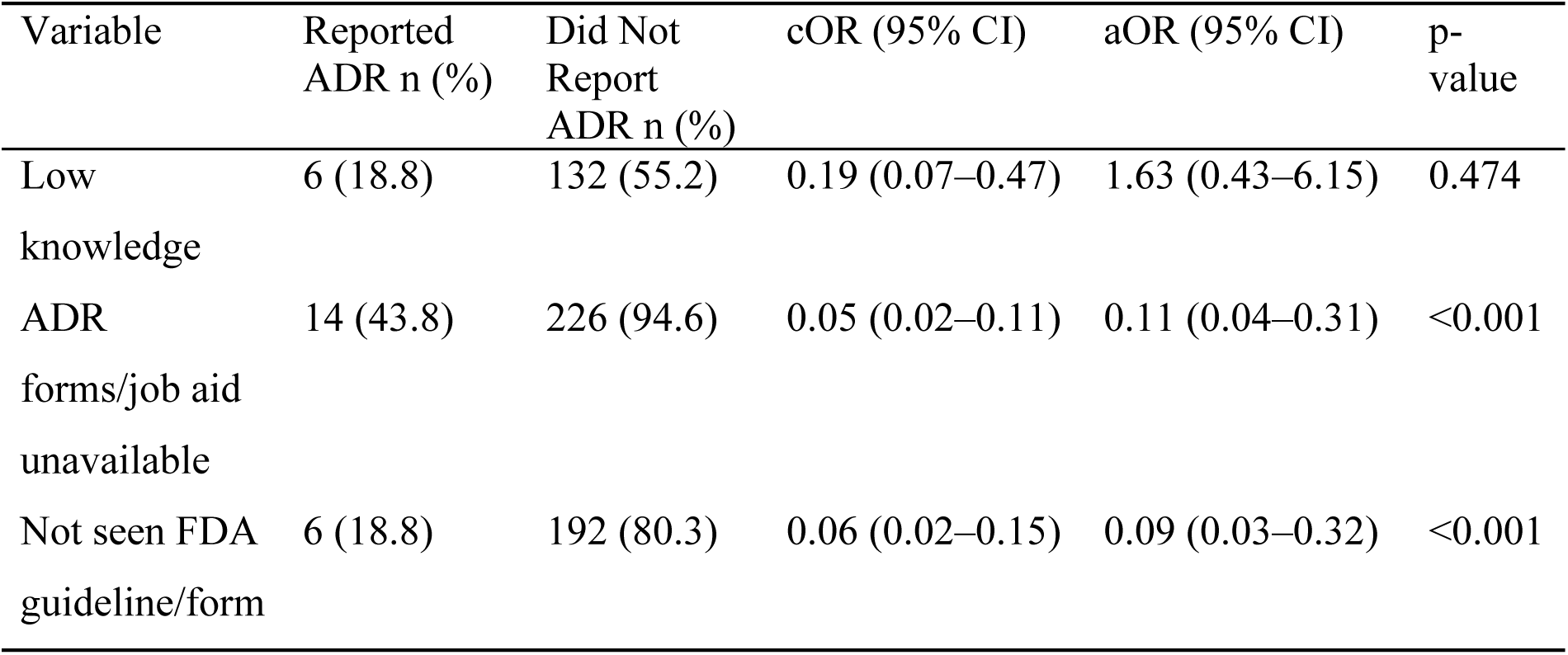
Multivariable Logistic Regression Analysis of Factors Associated with ADR Reporting Among Herbal Practitioners.

In the multivariable logistic regression model, system-level factors remained independently associated with ADR reporting. Practitioners working in facilities without ADR reporting forms or job aids had significantly reduced odds of reporting ADRs (aOR = 0.11, 95% CI: 0.04–0.31). Similarly, respondents who had not seen the Ghana FDA ADR reporting guideline or reporting form were less likely to report ADRs (aOR = 0.09, 95% CI: 0.03–0.32). ADR knowledge level was no longer independently associated with ADR reporting after adjustment for system-level factors.

### Qualitative Findings

The qualitative findings provided further insight into the barriers and facilitators of ADR reporting identified in the quantitative analysis and helped explain the broader contextual and systemic factors influencing reporting behaviour among herbal practitioners.

Two key themes emerged from the qualitative interviews: barriers to adverse drug reaction (ADR) reporting and strategies to improve ADR reporting. These themes reflected participants’ experiences with pharmacovigilance practices within herbal medicine settings and highlighted both structural and practitioner-level factors influencing ADR reporting behaviour

## Barriers to ADR Reporting

### Causal Uncertainty and Difficulty Attributing ADRs

A dominant barrier identified across interviews was the difficulty practitioners experienced in establishing clear causal relationships between herbal medicinal products and suspected adverse reactions. Participants explained that many patients were unable to identify the specific herbal product used, particularly where products were obtained from informal sources, self-prepared, or poorly labelled. This uncertainty was further complicated by concurrent use of multiple therapies, including conventional medicines.

One participant explained:

*“They come with complaints, but they do not even know which medicine they took, so it becomes very difficult to identify what exactly caused the reaction. You cannot even trace where it came from or what exactly they used.” **(Participant 2, Male, Herbal Medicine Practitioner)***

Several participants also described challenges associated with polypharmacy and simultaneous use of herbal and orthodox medicines, which complicated attribution of adverse reactions to a specific product:

*“When patients mix herbal medicine with other drugs, it becomes very difficult to know which one actually caused the adverse reaction, because you cannot separate the effects of each medicine.” **(Participant 9, Male, Herbal Medicine Practitioner)***

*“Sometimes the patient will just tell you that they took some herbal medicine, but they cannot mention the name or where they got it from. So, it becomes very difficult to link the reaction to a particular product.” **(Participant 4, Male, Herbal Doctor)***

*“In some cases, they are taking multiple things at the same time, including over-the-counter drugs, so even if there is a reaction, you are not sure whether it is from the herbal medicine or something else.” **(Participant 3, Male, Consulting Herbal Medicine Practitioner)***

### Inadequate Training and Clinical Assessment Gaps

Participants consistently highlighted inadequate pharmacovigilance training and limited clinical assessment skills as important barriers affecting ADR recognition and reporting. Although some participants demonstrated general awareness of adverse reactions, many described limited exposure to structured training on ADR identification and reporting procedures. A pharmacist explained:

*“At my workplace, we have not had structured training on adverse drug reaction reporting, so many people may not even know how to go about it or even where to start from.” **(Participant 1, Female, Pharmacist)***

Participants further indicated that inadequate clinical assessment skills affected practitioners’ ability to recognize ADRs during routine practice:

*“Some practitioners do not have the knowledge to even recognize that what they are seeing is an adverse drug reaction. They may just treat it as a normal condition and move on.” **(Participant 3, Male, Consulting Herbal Medicine Practitioner)***

*“If you are not trained to identify these reactions, you may not even notice them when they occur, and that means you will not think about reporting them at all.” **(Participant 11, Male, Herbal Medicine Practitioner)***

*“Training is very important because without it, people may not understand what qualifies as an adverse reaction, and that affects whether they report it or not.” **(Participant 12, Female, Community Herbal Retailer)***

### Weak Reporting Systems and Lack of Documentation

Weak reporting structures and poor documentation practices emerged as another barrier to ADR reporting. Participants described reporting processes within many herbal medicine settings as informal, fragmented, and poorly institutionalized. In many instances, adverse reactions were managed verbally without structured documentation or escalation to regulatory authorities. Participants shared that:

*“Most of the time, when people complain, it is handled verbally and there is no proper documentation of the reaction, so it ends there.” **(Participant 8, Female, Medicine Counter Assistant)***

Other participants emphasized the lack of clear reporting pathways and argued that the lack of documentation demotivates ADR reporting:

*“In some places, there is no proper structure for reporting, so people just talk about it among themselves without putting anything down officially.” **(Participant 6, Male, Community Herbal Retailer**)*

*“We do not have a clear system for reporting, so even if something happens, it may not be formally recorded.” **(Participant 10, Male, Medicine Counter Assistant)***

*“Even when cases are observed, there is no formal process to document and escalate it, so it becomes difficult to include it in any official reporting system.” **(Participant 2, Male, Herbal Medicine Practitioner)***

### Commercial Interests and Weak Regulation

Participants also identified commercial pressures and weak regulatory enforcement as factors discouraging ADR reporting. Some participants explained that practitioners involved in producing or selling herbal products may be reluctant to report adverse reactions because of concerns about reputational damage, reduced consumer trust, and potential financial losses. A male herbal doctor said:

*“If someone has invested money in producing a product, they may not want to report negative reactions because it will affect their business and reduce their sales.” **(Participant 5, Male, Herbal Doctor/Nutritionist)***

Similarly, another participant noted:

*Some people are more focused on the business side, so even if there is a problem with the product, they may not want to bring it out because it will affect their income.” **(Participant 6, Male, Community Herbal Retailer)***

*“The regulation is not strong enough, so people can practice without proper oversight, and that affects how adverse reactions are handled and reported.” **(Participant 7, Male, Traditional Herbalist)***

*“It shouldn’t be a free-for-all kind of thing where anybody can just do anything, because that makes it difficult to control what is happening in the system.” (**Participant 4, Male, Herbal Doctor)***

### Limited Government Prioritization of Herbal Medicine

Participants perceived that herbal medicine pharmacovigilance receives limited policy attention and institutional support. This perceived lack of prioritization was described as contributing to weak reporting structures, limited practitioner support, and inadequate investment in herbal medicine safety monitoring. One participant stated:

*“One of the challenges is that we see it to be that the government has no interest in herbal medicine, so there are no strong systems to support reporting.” **(Participant 2, Male, Herbal Medicine Practitioner)***

Others highlighted the absence of structured support systems and guidance from regulatory authorities:

*“There is not enough support from the authorities in terms of training and systems, so practitioners are left to handle things on their own.” **(Participant 5, Male, Herbal Doctor/Nutritionist)***

*“If the government gives more attention to herbal medicine and puts proper systems in place, it will improve how adverse reactions are reported.” **(Participant 12, Female, Community Herbal Retailer)***

*“If there were proper structures from the government, we would know exactly what to do and how to report these issues, but that is not the case now.” **(Participant 3, Male, Consulting Herbal Medicine Practitioner)***.

### Strategies to Improve ADR Reporting

Participants proposed several strategies to strengthen ADR reporting practices within herbal medicine settings. Suggested interventions focused on improving practitioner capacity, strengthening reporting infrastructure, enhancing regulatory oversight, and improving institutional support systems.

### Continuous Training and Sensitization

Participants consistently emphasized the need for continuous pharmacovigilance training and sensitization programmes to improve awareness, recognition, and reporting of ADRs. One participant explained:

*“Continuous training is very important because if practitioners do not really understand what adverse reactions are, they may not even recognize it when they see it. Sometimes people may think it is just part of the illness or something normal, but with proper training, you will be able to identify it and also know how to report it appropriately.” **(Participant 4, Male, Herbal Doctor)***

Participants further indicated that regular educational programmes could improve practitioners’ confidence and motivation to report ADRs:

*“Education and sensitization will help because when people understand the importance of reporting, they will take it seriously. Without that awareness, people may ignore these reactions or not see the need to report them.” **(Participant 12, Female, Community Herbal Retailer)**”*

*“There should be regular workshops and education for practitioners, not just once in a while. It should be something continuous so that people keep updating their knowledge, because if you don’t keep learning, you may forget or not even be aware of new reporting procedures that are being introduced.” **(Participant 3, Male, Consulting Herbal Medicine Practitioner)***

### Standardized and Accessible Reporting Systems

Participants also highlighted the need for simplified, user-friendly, and accessible reporting systems. Many believed that complicated reporting procedures discouraged practitioners from documenting and reporting ADRs. A pharmacist emphasized the importance of clarity and ease of use in reporting tools:

*“We need clear reporting forms and systems that are easy to use because if the process is complicated, people will not bother to report. It should be something straightforward that anyone can follow without difficulty, otherwise it becomes a burden and people will ignore it.” **(Participant 1, Female, Pharmacist)***

Other participants proposed accessible digital platforms and clearer reporting pathways:

*“If there is a simple system or platform that people can easily access, I believe more people will be willing to report. Right now, some people may not even know where to report or how to go about it, so they just leave it.” **(Participant 8, Female, Medicine Counter Assistant)***

*“Sometimes the problem is not that people do not want to report, but they do not know the process or the system is not clear. If the reporting system is made simple and accessible, it will encourage more reporting.” **(Participant 10, Male, Medicine Counter Assistant)***

*“If there is a proper structure where you know exactly who to report to and how to do it, it makes it easier. Without that structure, people may not take the extra step to report.” **(Participant 6, Male, Community Herbal Retailer)***

### Strengthened Regulation and Licensing

Participants further recommended strengthening regulatory and licensing systems to improve accountability and professional standards within herbal medicine practice. One participant explained:

*“There should be stricter regulations so that only trained professionals are allowed to practice, because when people are properly trained, they will understand the importance of reporting adverse reactions and also follow the right procedures.” **(Participant 2, Male, Herbal Medicine Practitioner)***

Other participants added:

*“Licensing should be tightened to make sure that only qualified people are involved in herbal medicine. If anyone can just enter the practice, then it becomes difficult to control how things are done, including how adverse reactions are handled.” **(Participant 5, Male, Herbal Doctor/Nutritionist)***

*“The system should not allow just anyone to practice because when untrained people are involved, they may not even understand the need to report adverse reactions. Regulation will help to control that.” **(Participant 7, Male, Traditional Herbalist)***

### Proactive Monitoring and Regulatory Supervision

Finally, participants emphasized the importance of proactive regulatory monitoring and supervision. Rather than relying solely on passive reporting after harm occurs, participants recommended routine inspections, monitoring, and early intervention by regulatory authorities. A male herbal doctor stated that:

*“Regulators should be more proactive rather than reactive. They should not wait for people to suffer or for complaints to come before they step in. There should be regular checks and monitoring so that potential problems can be identified early before they become serious.” **(Participant 4, Male, Herbal Doctor)***

Others argued that continuous supervision could improve practitioner accountability and reporting compliance:

*“Regular supervision will help practitioners follow proper procedures, because when people know that there is monitoring, they are more likely to do the right thing, including reporting adverse reactions when they occur.” **(Participant 6, Male, Community Herbal Retailer)***

“There should be continuous monitoring from the authorities so that practitioners are guided and corrected when necessary. This will improve how things are done, including reporting.” **(Participant 2, Male, Herbal Medicine Practitioner)**

“If the authorities are actively involved and checking what is happening on the ground, it will make practitioners more accountable and encourage proper reporting practices.” **(Participant 5, Male, Herbal Doctor/Nutritionist).**

Across the three study components, the findings converged on limited ADR reporting and important system-level barriers. The national pharmacovigilance data showed a relatively small number of herbal medicine-related ADR reports over the 2014–2024 period, while only 11.8% of surveyed herbal practitioners reported submitting at least one ADR report in the preceding 12 months. In the multivariable analysis, lack of access to ADR reporting forms or job aids and limited exposure to FDA reporting guidance were associated with lower odds of reporting. The qualitative findings provided contextual explanations for these patterns, with participants describing weak documentation systems, limited access to reporting tools and guidance, inadequate training, and uncertainty in attributing reactions to herbal medicinal products.

## Discussion

### Summary of Main Findings

This mixed-methods study assessed trends in herbal medicine-related adverse drug reaction (ADR) reporting in Ghana, determined the prevalence and factors associated with ADR reporting among herbal practitioners, and explored barriers affecting reporting practices. The findings demonstrated persistently low reporting of herbal medicine-related ADRs within the national pharmacovigilance system and low ADR reporting prevalence among herbal practitioners. Secondary pharmacovigilance data showed fluctuating but generally low numbers of herbal medicine-related ADR reports between 2014 and 2024, with reporting concentrated largely in the Greater Accra Region. In the quantitative component, only 11.8% of herbal practitioners reported submitting an ADR report within the previous 12 months, highlighting low ADR reporting within the herbal medicine sector.

The study further demonstrated that ADR reporting behaviour was associated more strongly with system-level factors than by individual practitioner characteristics. Lack of ADR reporting forms or job aids and lack of exposure to Ghana Food and Drugs Authority (FDA) reporting guidelines was associated with significantly lower odds of ADR reporting. Although ADR knowledge and formal training were associated with reporting at the bivariate level, these associations lost significance after adjustment for system-level factors, suggesting that institutional readiness and access to reporting infrastructure may play a more central role in influencing reporting behaviour. These findings align with evidence reported in previous pharmacovigilance studies, where inadequate reporting systems, limited institutional support, and poor access to reporting tools were identified as major barriers to ADR reporting^5, 6, 14^.

The qualitative findings provided contextual explanations for the quantitative findings by highlighting uncertainty in attributing ADRs to herbal medicines, inadequate pharmacovigilance training, weak documentation systems, commercial concerns, and limited regulatory engagement as major barriers affecting reporting practices. Collectively, the results indicate that underreporting of herbal medicine-related ADRs in Ghana is largely associated with structural constraints rather than inadequate practitioner knowledge alone.

### Low Prevalence of ADR Reporting

The 12-month prevalence of ADR reporting among herbal practitioners was low, with only 11.8% of respondents reporting submission of at least one ADR report within the previous year. This finding reflects substantial underreporting within the herbal medicine sector and is consistent with studies conducted among healthcare professionals in LMICs where ADR reporting rates remain suboptimal despite increasing pharmacovigilance awareness^6, 13, 15^. The identification of 117 eligible herbal medicine-related ADR reports over a ten-year period despite the long observation period, is consistent with substantial underreporting of herbal medicine-related ADRs within the formal pharmacovigilance system. However, spontaneous reporting data cannot be used to estimate the incidence or true burden of ADRs.

The low reporting prevalence may partly reflect the complexities associated with herbal medicine pharmacovigilance. Herbal medicinal products often have variable composition, limited standardization, inadequate labelling, and are frequently used concurrently with conventional medicines, making causality assessment difficult^16, 17^. Supporting this finding, interview narratives revealed difficulties linking suspected reactions to specific herbal products because of poor patient recall, polypharmacy, and unregulated product sourcing.

Underreporting remains a major limitation of spontaneous pharmacovigilance systems, particularly in resource-limited settings where reporting systems are poorly institutionalized^18^. Similar barriers, including limited reporting tools, uncertainty regarding reporting procedures, and weak institutional support, have been reported in other African settings^19, 20^. Persistent underreporting may weaken post-market surveillance, delay detection of safety signals, and underestimate the true burden of herbal medicine-related adverse events. Strengthening ADR reporting among herbal practitioners is therefore essential for improving herbal medicine safety surveillance in Ghana.

### Weak Pharmacovigilance Infrastructure and System-Level Barriers

One of the most important findings was the strong influence of structural barriers on ADR reporting behaviour among herbal practitioners. Lack of ADR reporting forms or job aids and lack of exposure to FDA reporting guidelines remained independently associated with reduced odds of ADR reporting after adjustment for individual-level factors. These findings suggest that institutional and structural barriers may exert greater influence on reporting practices than practitioner knowledge alone.

The availability of reporting forms and job aids was one of the strongest predictors of ADR reporting. Comparable observations have been documented in previous studies where inadequate reporting infrastructure and limited access to reporting tools reduced pharmacovigilance reporting practices^19–21^. Exposure to reporting guidelines may also improve familiarity with reporting procedures and increase confidence in identifying and reporting suspected ADRs.

Interview data revealed weak reporting structures, poor documentation practices, and limited institutional support within herbal medicine settings. Several participants described ADR complaints being managed informally without proper documentation or escalation to regulatory authorities. These findings suggest that herbal medicine pharmacovigilance in Ghana remains weakly institutionalized and largely dependent on passive and inconsistent reporting practices.

An important observation was that ADR knowledge and formal training lost significance after adjustment for system-level variables. This suggests that although knowledge is important, it may not translate into reporting behaviour in the absence of supportive institutional systems and accessible reporting infrastructure. Similar findings have been reported in previous pharmacovigilance studies where healthcare professionals demonstrated adequate awareness but still exhibited poor reporting practices due to weak reporting systems and limited institutional support^7, 22^.

These findings suggest that improving ADR reporting within the herbal medicine sector may require broader structural reforms rather than awareness creation alone. Strengthening institutional pharmacovigilance structures, increasing access to reporting tools, improving dissemination of FDA guidelines, and integrating herbal practitioners more effectively into national pharmacovigilance systems may substantially improve herbal medicine safety surveillance in Ghana.

### Knowledge and Training Issues

Although knowledge and training were associated with ADR reporting at the bivariate level, these associations disappeared after adjustment for system-level factors, suggesting that reporting behaviour is more strongly linked to the availability of reporting infrastructure and institutional support than to individual practitioner characteristics. Nevertheless, the observed bivariate associations suggest that knowledge and training remain important for effective engagement with ADR reporting systems. Participants with higher ADR knowledge were more likely to report ADRs during the initial analysis, consistent with previous studies among healthcare professionals^6, 13^. Similarly, formal training may improve practitioners’ ability to identify and document suspected adverse reactions. However, the attenuation of these associations after adjustment suggests an important interaction between individual knowledge and institutional systems.

The qualitative findings highlighted important gaps in pharmacovigilance literacy among herbal practitioners. Several participants reported limited exposure to structured ADR reporting training and uncertainty regarding reportable ADRs and reporting procedures. These findings reflect broader challenges within herbal medicine pharmacovigilance, where training opportunities for herbal practitioners may be limited compared with conventional healthcare professionals.

The findings therefore suggest that interventions aimed at improving reporting practices should extend beyond awareness creation and incorporate simplified reporting procedures, stronger Institutional support, and regular engagement between the Ghana FDA and herbal practitioners.

### Qualitative Interpretation of Underreporting

The interview data provided important contextual explanations for the persistently low ADR reporting observed across the secondary and quantitative data. Participants described uncertainty in attributing ADRs to herbal medicines, weak documentation practices, commercial concerns, and limited institutional support as major contributors to underreporting.

Causal uncertainty emerged as a particularly important barrier. Participants described difficulties determining whether adverse reactions were directly caused by herbal medicines, especially where patients used multiple therapies simultaneously or obtained products from unregulated sources. These findings are consistent with earlier reports in which uncertainty regarding causality reduced confidence in reporting among healthcare professionals^7, 8^.

Weak documentation practices also contributed substantially to underreporting. Several participants reported that ADR complaints were often managed informally without structured documentation or reporting mechanisms. These findings are consistent with previous studies conducted in low-resource settings where inadequate documentation systems negatively affected pharmacovigilance practices^16, 23^.

Commercial and reputational concerns were also identified as barriers influencing reporting behaviour. Some participants expressed concern that reporting adverse reactions associated with herbal products could negatively affect product sales and consumer trust. Similar concerns regarding reputational damage and fear of blame have been reported in previous pharmacovigilance studies ^8, 9^.

### Implications for Pharmacovigilance Policy and Practice

The findings have implications for strengthening herbal medicine pharmacovigilance in Ghana. Improving access to standardised ADR reporting tools, simplifying reporting procedures, and routinely disseminating FDA reporting guidance could address key system-level barriers identified in the study. Targeted pharmacovigilance training should complement these measssures, particularly where practitioners have limited exposure to reporting procedures. Strengthening documentation systems, establishing regular feedback mechanisms between regulatory authorities and herbal practitioners, and improving post-market surveillance and regulatory monitoring may further support reporting, transparency, accountability, and product traceability. Greater collaboration between the Ghana FDA and the Traditional Medicine Practice Council could support more consistent engagement of herbal practitioners in the national pharmacovigilance system. Addressing commercial concerns through supportive regulatory oversight and strengthening trust between practitioners and regulatory authorities may also reduce reluctance to report suspected ADRs.

### Strengths and Limitations

This study has several important strengths. The mixed-methods design enabled triangulation of secondary pharmacovigilance data with quantitative and qualitative findings, thereby strengthening the credibility and depth of the findings. The inclusion of national pharmacovigilance data from the Ghana FDA provided important contextual information on herbal medicine-related ADR reporting trends over a ten-year period. In addition, the use of probability sampling and multivariable logistic regression improved the analytical rigor of the quantitative component, while qualitative interviews provided contextual explanations for the observed reporting patterns.

However, some limitations should be considered. The cross-sectional design limits causal inference between explanatory variables and ADR reporting behaviour. The quantitative findings were also based on self-reported practices and may therefore be affected by recall and social desirability bias. In addition, the secondary pharmacovigilance data may underestimate the true burden of herbal medicine-related ADRs because spontaneous reporting systems are inherently affected by underreporting.

Furthermore, the study was conducted among registered herbal practitioners within selected districts of the Greater Accra Region and may not fully represent ADR reporting practices among unregistered practitioners or those operating in other regions of Ghana, particularly rural settings where herbal medicine practices and reporting systems may differ.

Nevertheless, the study provides important evidence on herbal medicine pharmacovigilance and contributes to the limited literature on ADR reporting among herbal practitioners in Ghana and similar low-and middle-income country settings.

## Conclusion

This mixed-methods study found that adverse drug reaction (ADR) reporting associated with herbal medicinal products was limited within the national pharmacovigilance system and among herbal practitioners surveyed in selected districts of the Greater Accra Region. The study demonstrated that underreporting is associated more strongly with system-level and institutional barriers than by individual practitioner characteristics alone. Lack of ADR reporting forms or job aids, limited exposure to Ghana Food and Drugs Authority (FDA) reporting guidelines, weak documentation systems, and inadequate institutional support were identified as major factors limiting ADR reporting practices among herbal practitioners.

The qualitative accounts reinforced important challenges affecting herbal medicine pharmacovigilance, including uncertainty in attributing ADRs to herbal medicinal products, inadequate pharmacovigilance training, poor documentation practices, commercial concerns, and limited regulatory engagement. Together, the findings suggest that herbal medicine pharmacovigilance in Ghana remains weakly institutionalized despite the widespread use of herbal medicinal products.

Strengthening herbal medicine pharmacovigilance systems in Ghana will require comprehensive system-level interventions that go beyond awareness creation alone. Improving access to reporting tools, increasing dissemination of ADR reporting guidelines, strengthening practitioner training, enhancing regulatory engagement, and integrating herbal practitioners more effectively into national pharmacovigilance systems may improve ADR reporting practices and medicine safety surveillance. Further research is needed to evaluate the effectiveness of interventions aimed at strengthening ADR reporting among herbal practitioners and to explore reporting practices in other regions of Ghana.

The study highlights the need for stronger institutional integration of herbal medicine practitioners into national pharmacovigilance systems in Ghana.

## Data Availability

The datasets generated and/or analysed during the current study are available from the corresponding author upon reasonable request.

## Abbreviations

ADR: Adverse Drug Reaction
aOR: Adjusted Odds Ratio
CI: Confidence Interval
FDA: Food and Drugs Authority
LMICs: Low-and Middle-Income Countries
MAXQDA: Max Qualitative Data Analysis Software
NMIMR: Noguchi Memorial Institute for Medical Research
SOC: System Organ Class
SPSS: Statistical Package for the Social Sciences
TMPC: Traditional Medicine Practice Council
WHO: World Health Organization

## Declarations

### Ethics Approval and Consent to Participate

Ethical approval for this study was obtained from the Noguchi Memorial Institute for Medical Research Institutional Review Board (NMIMR-IRB CPN 020/25-26). Permission to conduct the study was obtained from relevant regulatory authorities and participating facilities prior to data collection. Written informed consent was obtained from all participants before participation in the study. Confidentiality and anonymity of participants were maintained throughout the study.

### Consent for Publication

Not applicable.

### Clinical trial number

Not applicable

### Authors’ Contributions

**HF** developed the research questions, study objectives, and methodology; conducted data collection; performed quantitative data analysis; interpreted the findings; and drafted the manuscript. **HB** conceptualized the study, supervised the research, contributed to study design and interpretation of the findings, and critically revised the manuscript for important intellectual content. **JS** contributed substantially to qualitative data analysis and interpretation, critically reviewed the manuscript, and assisted with reference management. **CS** contributed to the qualitative component of the study, including interpretation of qualitative findings, and critically revised the manuscript for important intellectual content. **FZ** critically reviewed the manuscript, contributed to improving its scientific quality and presentation, and provided substantial intellectual input during manuscript revision. **GTS** provided technical guidance on the pharmacovigilance component and critically reviewed the manuscript for intellectual content. **KFO, SKS, and PB** provided critical review and substantive intellectual input to the manuscript and contributed to its revision. All authors read and approved the final manuscript and agree to be accountable for all aspects of the work

## Acknowledgements

The authors are grateful to the Ghana Food and Drugs Authority (FDA) for its institutional support and for granting access to the national pharmacovigilance data used in this study. We thank the Traditional Medicine Practice Council (TMPC) for providing the sampling frame of registered herbal practitioners and all herbal practitioners who participated in this study. We also acknowledge all research assistants for their support during data collection, with special appreciation to Lawrencia Abrafi Osei for her valuable contribution.

---

**Table 1.**
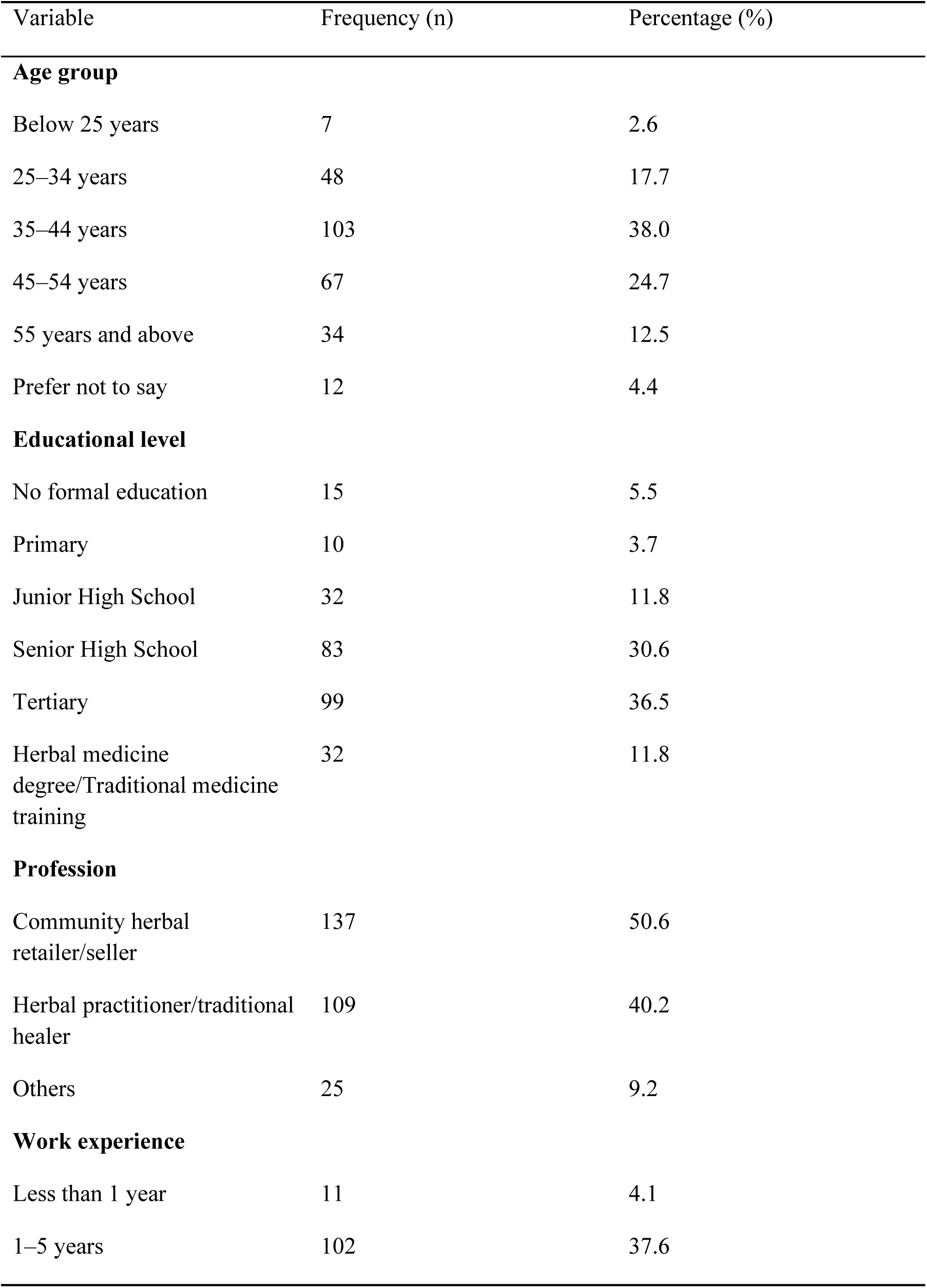
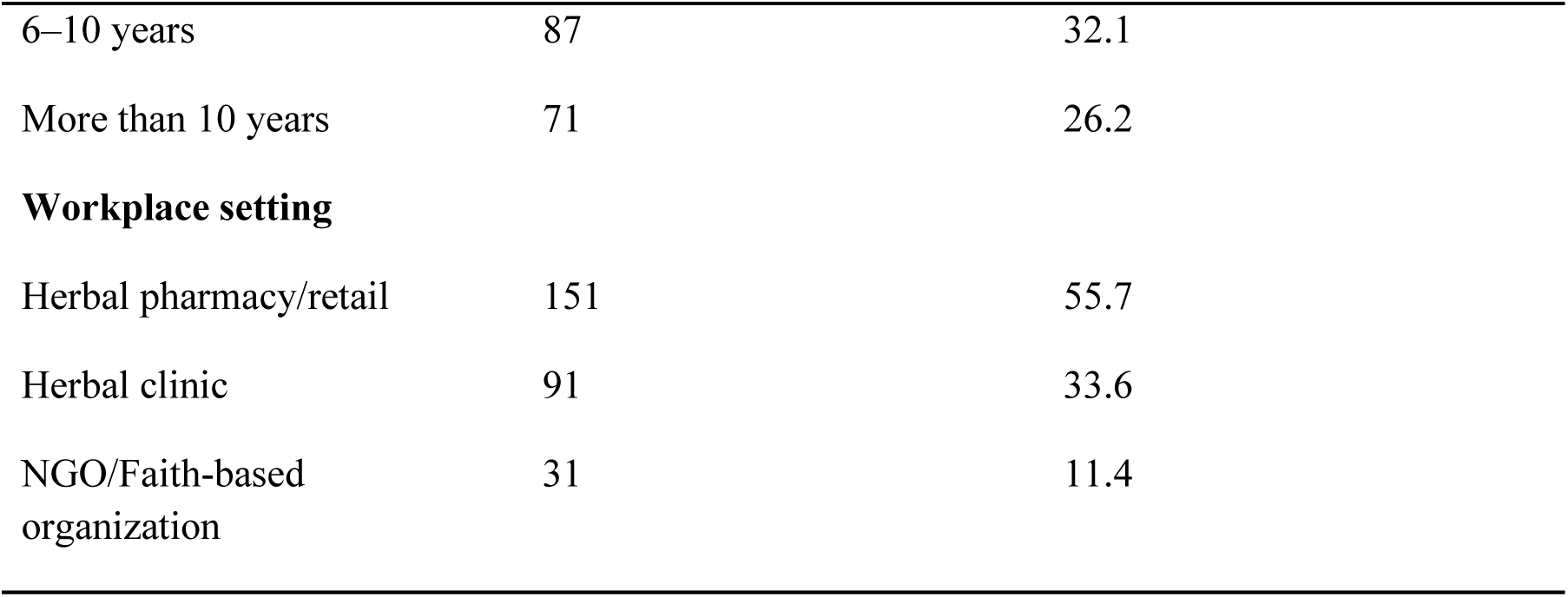
Socio-demographic Characteristics of Herbal Practitioners Participating in the Study (n = 271)

**Table 2.**
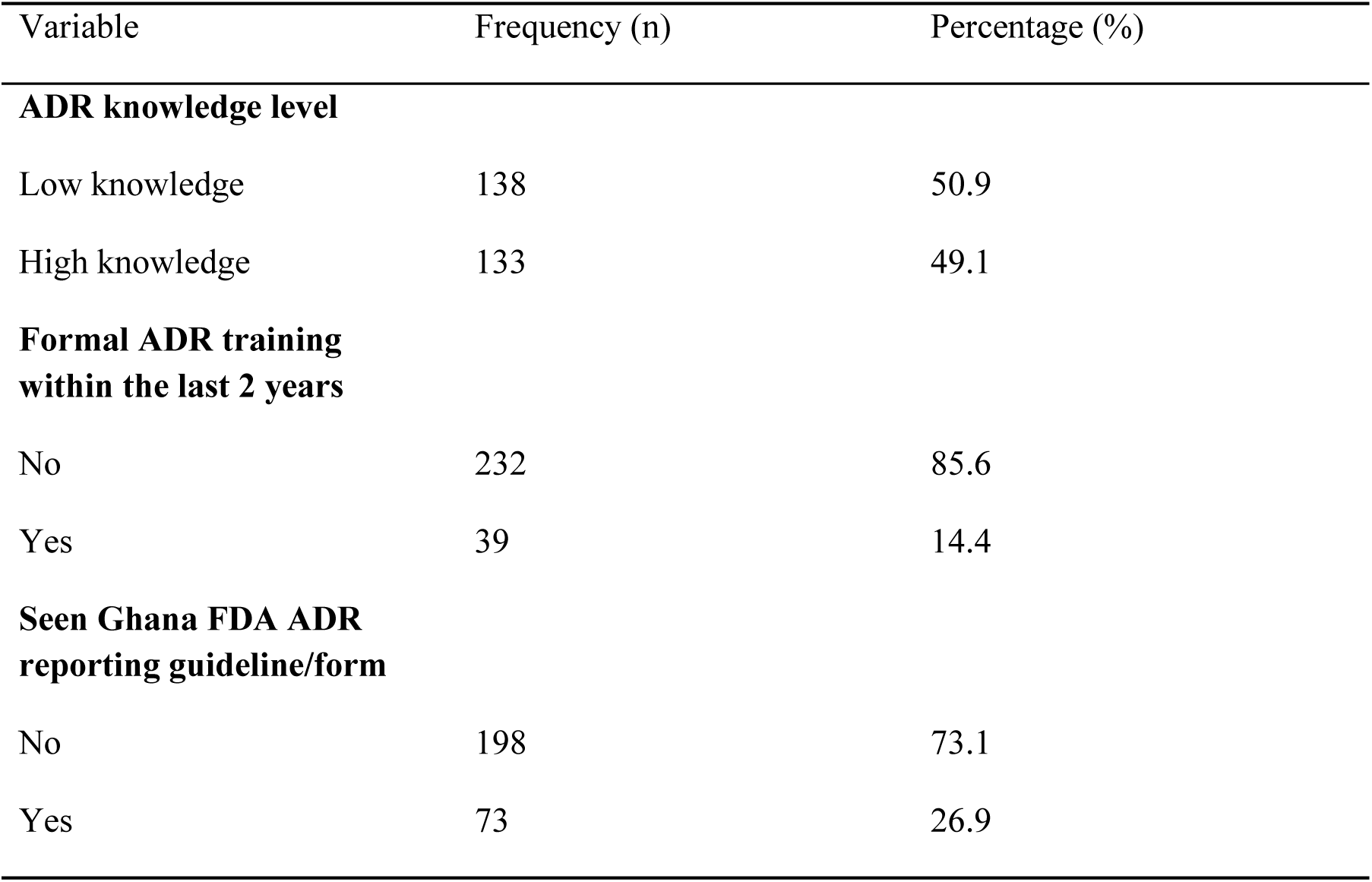
Knowledge, Training, and System-Related Characteristics of Herbal Practitioners.

**Table 3.**
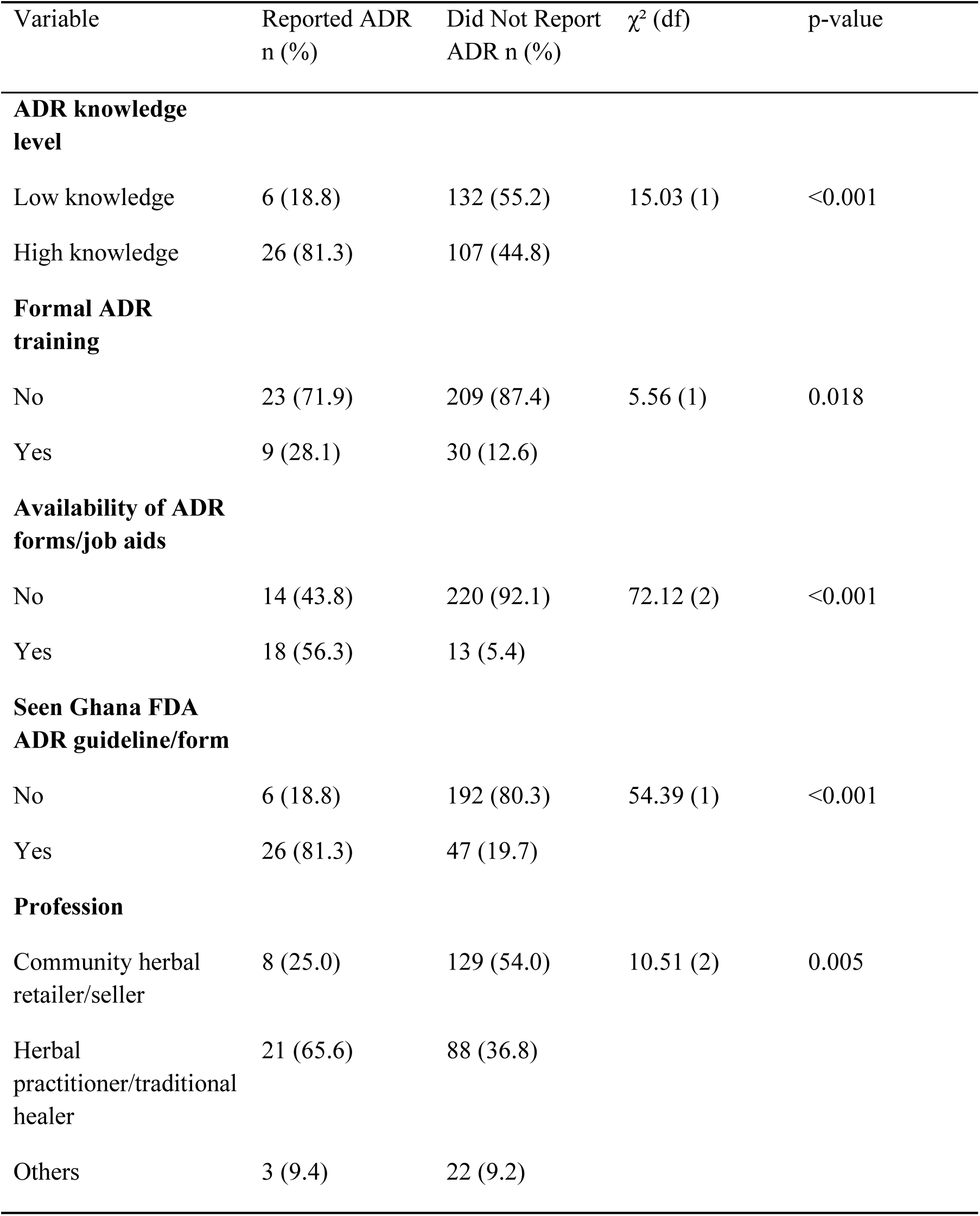
Bivariate Analysis of Factors Associated with ADR Reporting Among Herbal Practitioners.

**Table 4.**
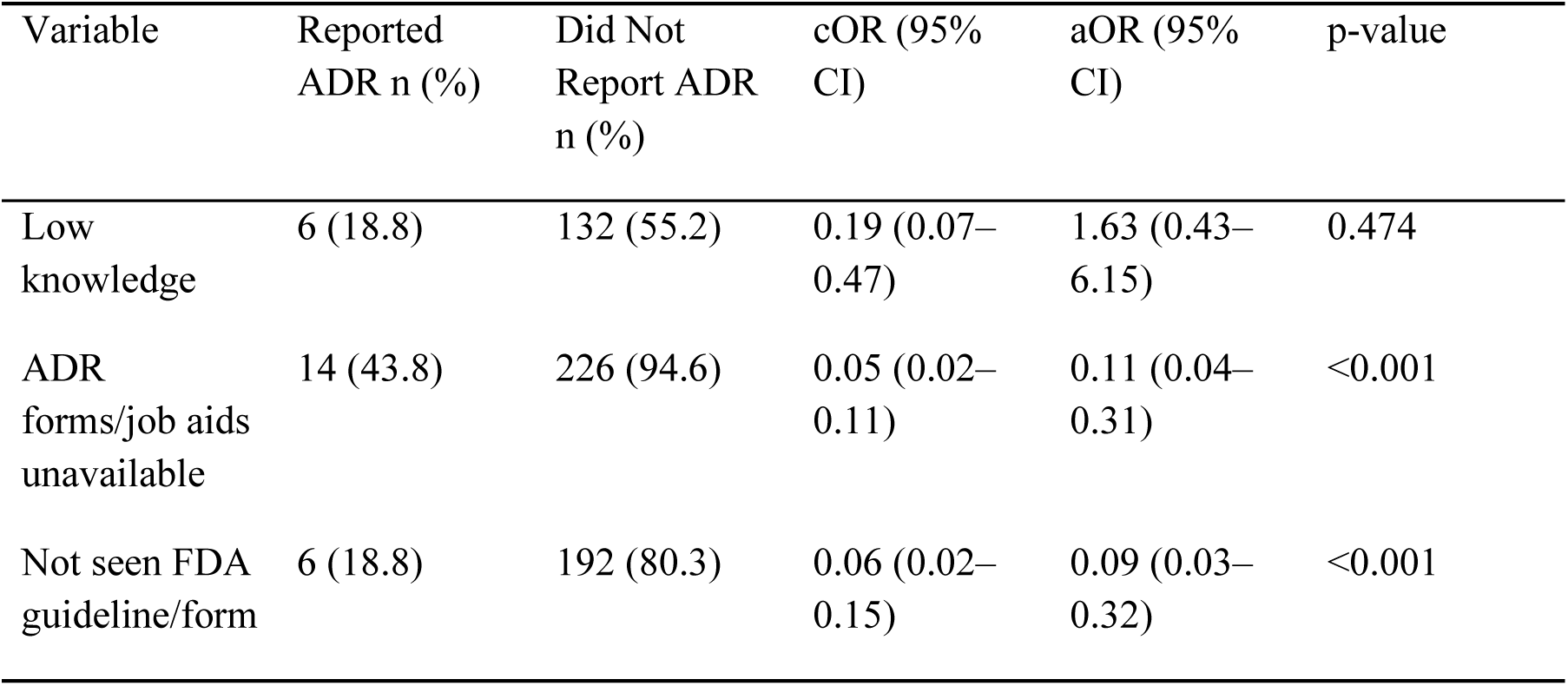
Multivariable Logistic Regression Analysis of Factors Associated with ADR Reporting Among Herbal Practitioners.

**Table 5.** Summary of Themes and Subthemes from Qualitative Interviews on Barriers and Strategies Related to ADR Reporting.

| Major Theme | Subtheme | Illustrative Quote |
| --- | --- | --- |
| Barriers to ADR Reporting | Causal uncertainty | “Patients often do not know exactly which medicine caused the reaction, making it difficult to confidently attribute the ADR.” |
| Barriers to ADR Reporting | Weak reporting systems | “Most complaints are handled verbally without proper documentation or reporting to the authorities.” |
| Barriers to ADR Reporting | Inadequate pharmacovigilance training | “At our facility, there has not been any structured ADR reporting training.” |
| Strategies to Improve ADR Reporting | Continuous training | “Continuous training is important because practitioners need to recognize ADRs and understand reporting procedures.” |

**Table 6.**
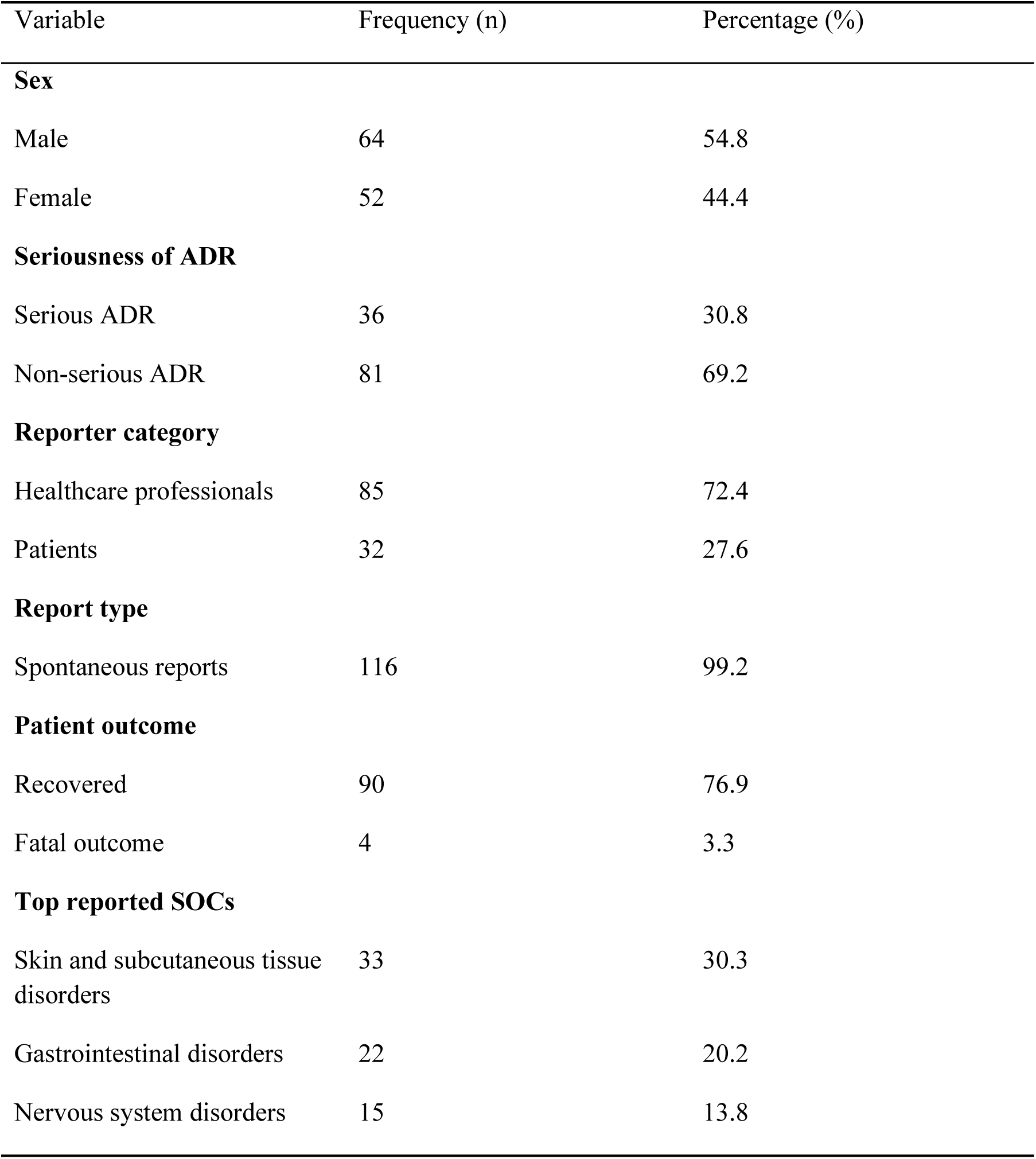
Characteristics of Herbal Medicine-Related ADR Reports Submitted to the Ghana FDA, 2014–2024 (n = 117)

